# Interethnic Validation of Artificial Intelligence for prediction of Atrial Fibrillation Using Sinus Rhythm Electrocardiogram

**DOI:** 10.1101/2025.03.09.25323529

**Authors:** Ji Hyun Lee, Joonghee Kim, Jina Choi, Yun Young Choi, Il-Young Oh, Youngjin Cho

## Abstract

**Background:** Previous research has demonstrated acceptable diagnostic accuracy of AI-enabled sinus rhythm (SR) electrocardiogram (ECG) interpretation for predicting paroxysmal or incident atrial fibrillation (AF). However, interethnic validations of these AI algorithms remain limited. We aimed to develop and comprehensively evaluate our AI model for predicting AF based on standard 12Dlead SR ECG images in a Korean population, and to validate its performance in Brazilian patient cohorts.

**Methods:** We developed a modified convolutional neural network model using a dataset comprising 811,542 ECGs from 121,600 patients at Seoul National University Bundang Hospital (2003–2020). Ninety percent of the patients were allocated to the training dataset, while the remaining 10% to the internal validation dataset. The model outputs a risk score (from 0 to 1) indicating the probability of concurrent paroxysmal or incident AF within 2 years, using standard-format 12Dlead SR ECG images. External validation was performed using the CODE 15% dataset, an open ECG dataset from the Telehealth Network of Minas Gerais, Brazil, by applying a 1:4 (AF:Non-AF) random sampling strategy.

**Results:** In the internal validation, our AI model achieved an Area Under the Receiver Operating Characteristic Curve (AUROC) of 0.907 (95% CI: 0.897-0.916), with a sensitivity of 80.6% and a specificity of 85.0% for AF prediction. Subgroup analyses showed an AUROC of 0.874 (95% CI: 0.856-0.891) for patients in routine health checkups or outpatient settings, and 0.852 (95% CI: 0.824-0.880) for patients with "Normal ECG" interpretations. In the external interethnic validation with the CODE 15% dataset, the AI model exhibited an AUROC of 0.884 (95% CI: 0.869-0.900), which increased to 0.906 (95% CI: 0.893-0.919) when adjusted for age and sex. In the subset of patients with "Normal ECG" interpretations, the AUROC was 0.826 (95% CI: 0.769-0.883), increasing to 0.861 (95% CI: 0.814-0.908) after applying the same adjustments.

**Conclusions:** Our AI-powered SR ECG interpretation model demonstrated excellent performance in predicting paroxysmal or incident AF, with valid performance in the Brazilian population as well. This suggests that the model has potential for broad application across different ethnic groups.

## Introduction

Cardiovascular diseases are a leading cause of morbidity and mortality worldwide, highlighting the need for comprehensive cardiac evaluations.^1^ In particular, atrial fibrillation (AF) poses a significant burden on healthcare systems due to its strong association with stroke, heart failure, and increased mortality.^2^ However, detecting AF is challenging because its episodes are often intermittent and asymptomatic, frequently evading conventional screening methods.^2^

Recent advances in artificial intelligence (AI) have transformed cardiac diagnostics.^3^ Several research teams have demonstrated that AI models can accurately predict AF risk from standard 12-lead electrocardiograms (ECGs) recorded during sinus rhythm (SR).^4–6^ These models enable the identification of patients at a risk for concurrent paroxysmal or future AF, facilitating targeted and cost-effective screening of high-risk populations. Nevertheless, most existing approaches rely on one-dimensional raw ECG signals, whereas in routine clinical practice, ECGs are typically stored and interpreted as images. This discrepancy poses practical challenges, particularly in telemedicine settings where ECG images are often obtained by scanning printed reports or photographing them.

Building on our previous work in developing an AI system for rapid emergency cardiac evaluation, we have expanded our efforts by creating ECG Buddy for Cardiologist (EBC), a comprehensive mobile application for cardiac health assessment.^7–10^ As EBC is designed to evaluate multiple facets of cardiac function,^7–10^ we have recently developed a new module aimed at predicting concurrent paroxysmal AF or incident AF using standard SR ECG images. Furthermore, to enhance its broader generalizability, an interethnic validation test was conducted, as EBC was originally developed using data from a Korean population.

In this study, we present a comprehensive evaluation of EBC’s paroxysmal and incident AF prediction module—including both internal and interethnic external validations. By demonstrating its reliable identification of AF risk in both Korean and international patient cohorts, we aim to establish EBC as a tool that facilitate earlier detection and contribute to more effective and timely management of AF.

## Methods

### Study Design

In this retrospective, observational study, we evaluated the performance of EBC in predicting AF using standard 12Dlead ECG images. The model generates a risk score indicating the probability of concurrent paroxysmal or incident AF within 2 years and was validated using both internal and external datasets. This study was approved by the Institutional Review Board of Seoul National University Bundang Hospital (IRB No.: B-2205-757-002), and informed consent was waived due to the retrospective study design.

### Development Dataset

ECG data were obtained from Seoul National University Bundang Hospital between 2003 to 2020 (Fig. 1). A total of 811,542 ECGs from 121,600 patients aged 18 years or older were screened. ECGs with insufficient quality—determined by an inDhouse algorithm—and those from patients with electronic pacemakers or a history of loop recorder usage were excluded. Ninety percent of the patients were randomly assigned to the training dataset, and the remaining 10% were allocated to the internal validation dataset (Fig. 1). Within the dataset, ECGs were further categorized based on their temporal relationship to an AF diagnosis. The AF group included SR ECG images recorded within two years prior to an initial AF diagnosis, totaling 284,531 ECGs. In contrast, the Non-AF group consisted of SR ECGs recorded more than two years before an initial AF diagnosis or from patients with no AF diagnosis, comprising 442,931 ECGs.

**Figure 1.**
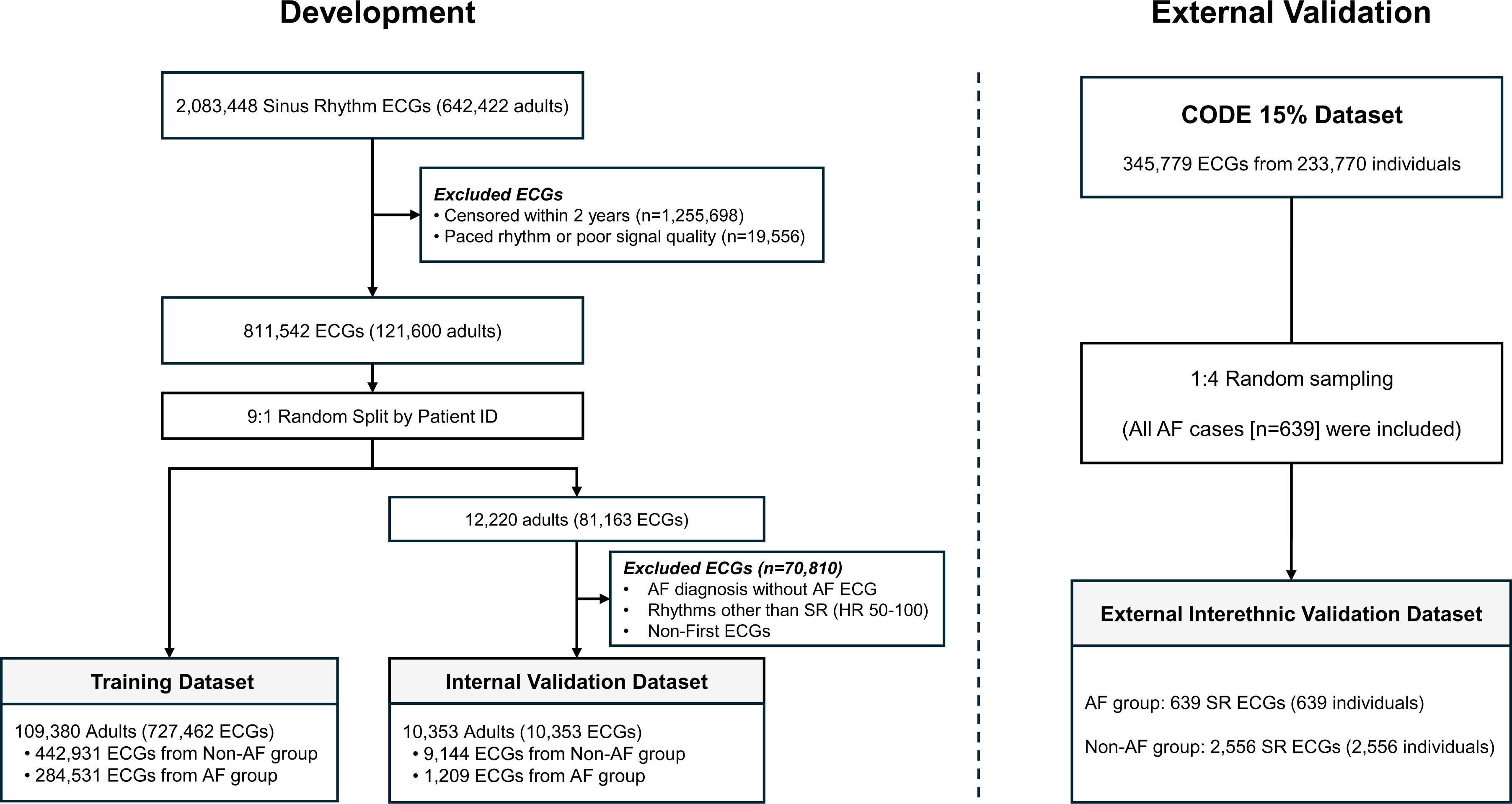
Flowchart illustrating the AI model development and external interethnic validation process. AF, atrial fibrillation; ECG, electrocardiogram; SR, sinus rhythm

### The Deep-Learning Model

The AF prediction component of EBC is built upon a modified convolutional neural network (CNN) architecture. The model consists of an encoder based on a ResNet backbone, enhanced with squeezeDexcitation layers and nonlocal blocks, and a taskDspecific module that outputs a risk score ranging from 0 to 1 for concurrent or incident AF within 2 years (AI-AF score). The encoder was pretrained using selfDsupervised learning schemes on the training dataset to extract robust numerical feature vectors from the input ECG images. These features were then fed into a multilayer perceptron, which produced the final risk score for paroxysmal or incident AF. StandardDformat images of 12Dlead ECGs (3×4 format with one rhythm strip [lead II]) as PNG image files were used as input data.

### Internal Validation Using the Test Dataset

The model’s ability to predict AF was evaluated on an internal test dataset comprising 81,163 ECGs from 12,220 patients, derived from the remaining 10% of the development dataset after applying additional exclusion criteria. Specifically, 1) ECGs from patients with an AF diagnosis without confirmation via ECG or Holter monitoring, and 2) rhythms other than Sinus Rhythm (SR) with a normal heart rate (50≤HR<100 bpm) were excluded. In internal validation, the AF group included the first SR ECG taken up to one month before the initial AF diagnosis (1,209 ECGs from 1,209 patients); the Non-AF group included the first SR ECG of patients with no AF diagnosis (9,144 ECGs from 9,144 patients).

### External Validation

External validation was performed using the CODE 15% dataset, an open-source dataset containing 15% of patients randomly selected via stratified sampling from the CODE dataset.^11^ The CODE dataset was originally collected by the Telehealth Network of Minas Gerais (TNMG), Brazil, between 2010 to 2016. TNMG is a public telehealth system provides service to 811 out of 853 municipalities in Minas Gerais, Brazil.^11^ The CODE 15% dataset includes a total of 345,779 ECGs from 233,770 patients, along with basic demographic character and comorbidities.^12^ Regarding the ethnic composition of Minas Gerais as of 2022, the two major ethnic group were Mixed (46.8%) and White (41.1%), followed by Black (11.8%), Indigenous (0.2%) and Asian (0.2%).^13^

The definitions of AF and Non-AF group remained the same as in the internal validation, except for the following modification: 1) the initial AF event was defined exclusively using ECGs, 2) the maximal time window between sinus rhythm ECG and AF was determined by a one-year age difference in the dataset, as the exact ECG recording dates within a given year were not available. Consequently, the potential time difference between two ECGs ranges from 1 (31 Dec to 1 Jan of the next year) to 729 days (1 Jan to 31 Dec of the following year), and 3) random sampling was applied to Non-AF group to maintain a AF:Non-AF group ratio of 1:4, reducing computational time and cost while preserving statistical balance.

### Statistical Analysis

The AI model’s performance was evaluated using the area under the receiver operating characteristic curve (AUC-ROC). Sensitivity, specificity, positive predictive value (PPV), and negative predictive value (NPV) were calculated by binning the AI score based on the Youden index. Performance evaluations were also conducted in two subgroups defined in the internal validation dataset: 1) ECGs done in healthcare checkups or outpatient settings, and 2) ECGs labeled as “normal ECG”. In the external validation dataset, only one subgroup was defined, which was the “normal ECG” group, as the information on the specific settings in which the ECG were obtained was not available in the CODE 15% dataset. A p-value less than 0.05 was considered significant. All statistical analyses were performed using R version 4.1.0 (R Foundation for Statistical Computing, Vienna, Austria).

## Results

### Development dataset and Internal Validation

A total of 811,542 ECGs of 121,600 patients were used for model development (Table 1, Fig. 1). Significant demographic differences were observed between the Non-AF and AF groups in both the training and validation datasets. Notably, the Non-AF group comprised older (63 vs 72, *p*<0.001) and higher proportion of male patients (62.4% vs 56.1%, *p*<0.001), compared with AF group in the training dataset (Table 1). Similar trends were also noted in the internal validation dataset.

**Table 1.**
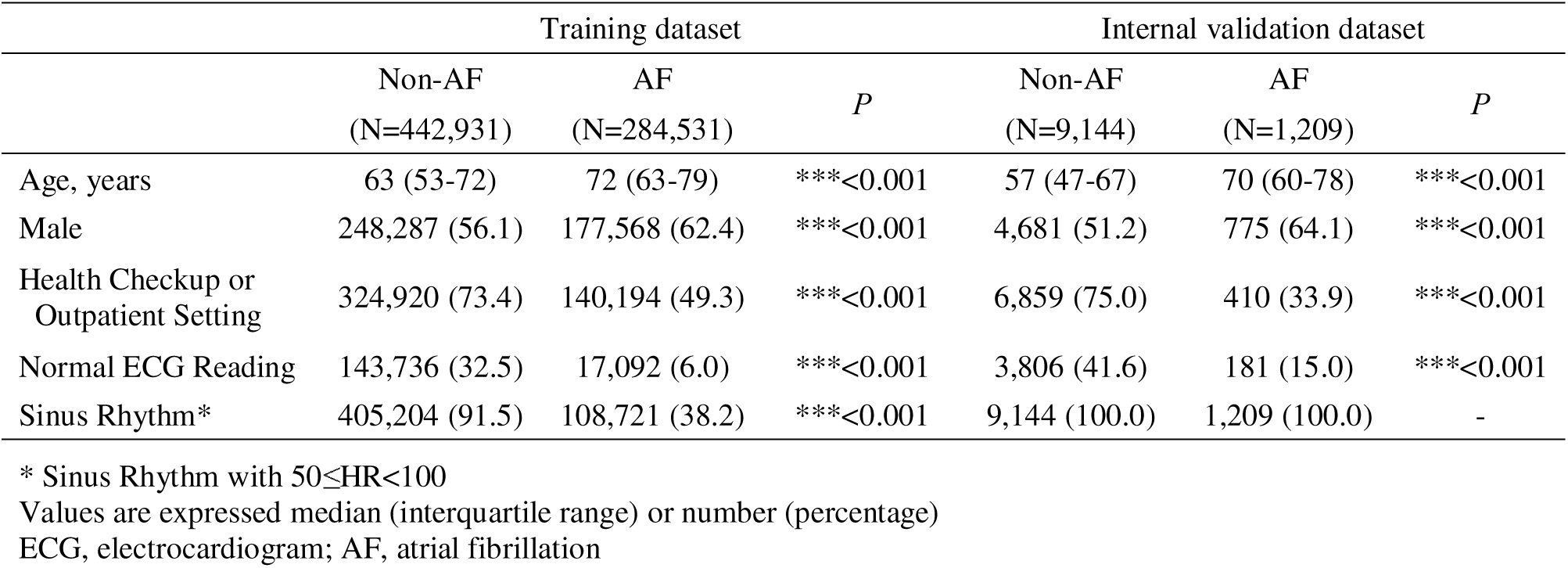
Description of the development dataset.

The internal validation of the AI model demonstrated robust performance in predicting AF, achieving an AUC-ROC of 0.907 (95% CI: 0.897-0.916), with a sensitivity of 80.6% (78.3-82.8%) and a specificity of 85.0% (84.2-85.7%) (Table 2). Additionally, the Positive Likelihood Ratio (PLR) was 5.36 (5.07-5.67) and the Negative Likelihood Ratio (NLR) was 0.23 (0.20-0.26). This high level of accuracy was consistently observed in subgroups, including ECGs from health checkup or outpatient visits settings (AUROC: 0.874) and those classified as “Normal ECG.” (AUROC: 0.852) (Table 2).

**Table 2.**
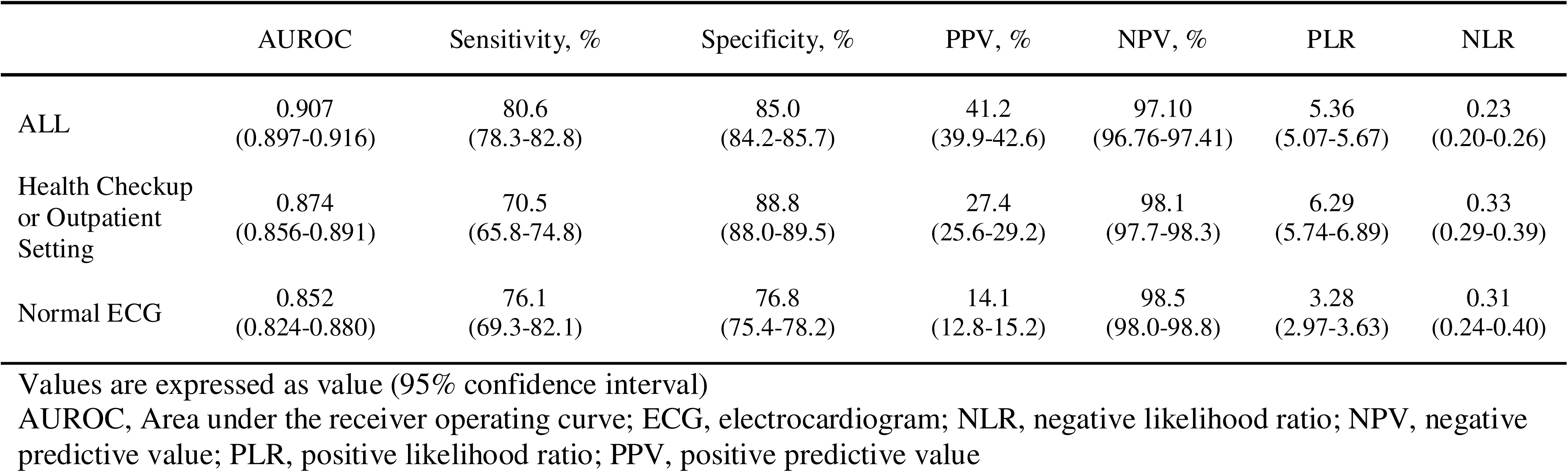
Internal validation performance.

### External Validation with CODE 15% dataset

External validation was conducted using 639 AF and 2,556 non-AF patients (Table 3) after 1:4 random sampling of the CODE 15% dataset. Similar to development datasets, significant demographic differences were noted between the Non-AF and AF groups, with AF patients being older (75 vs 50, *p*<0.001) and predominantly male (56.2% vs 41.6%, *p*<0.001) (Table 3). Figure 2 illustrates the distribution of AI-AF risk scores among AF and non-AF patients, showing significant difference (Median value: 0.754 vs. 0.133, *p*<0.001).

**Figure 2.**
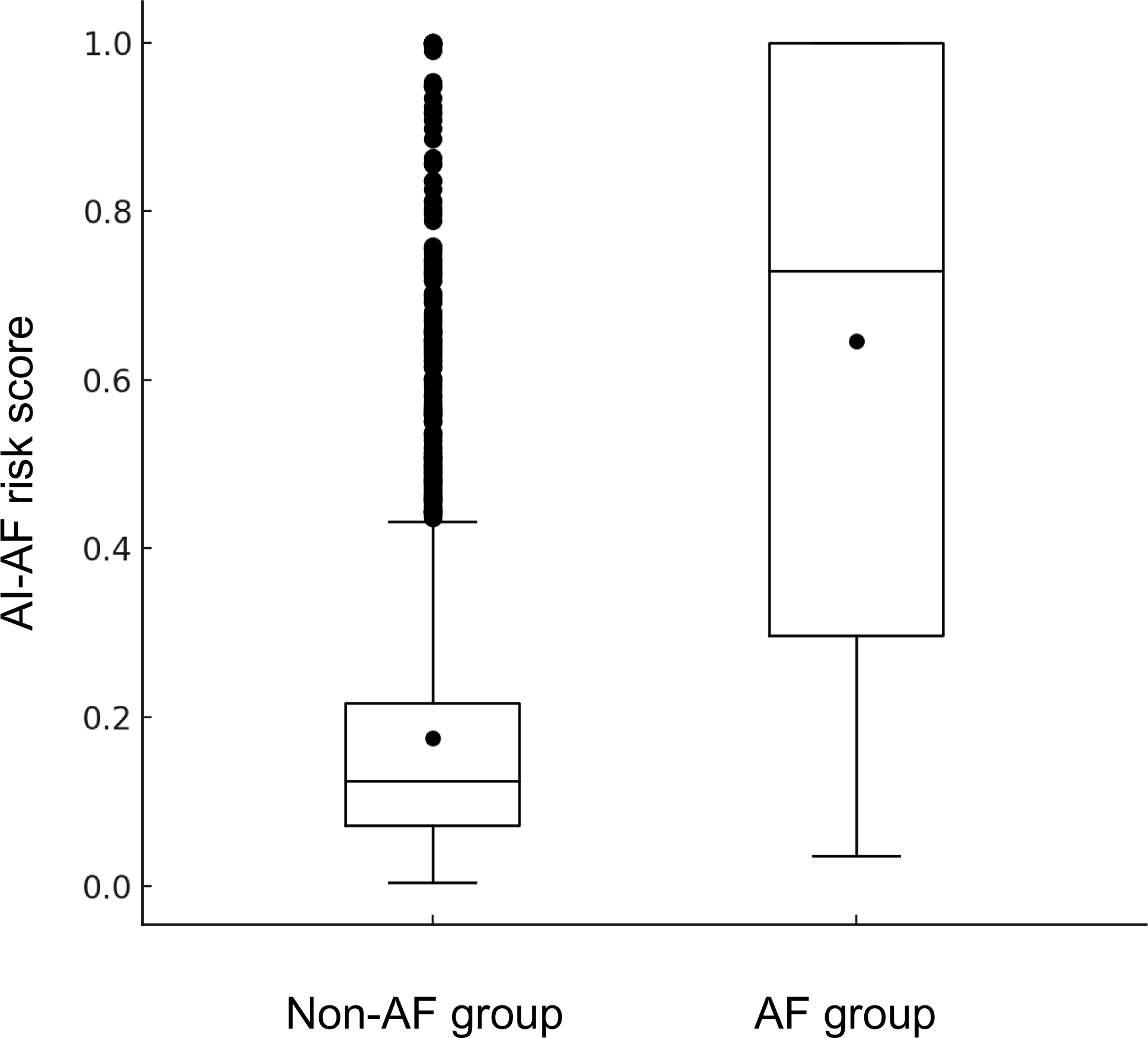
Box-plots, comparing the distribution of the AI-AF risk scores between study groups. AF, atrial fibrillation

**Table 3.**
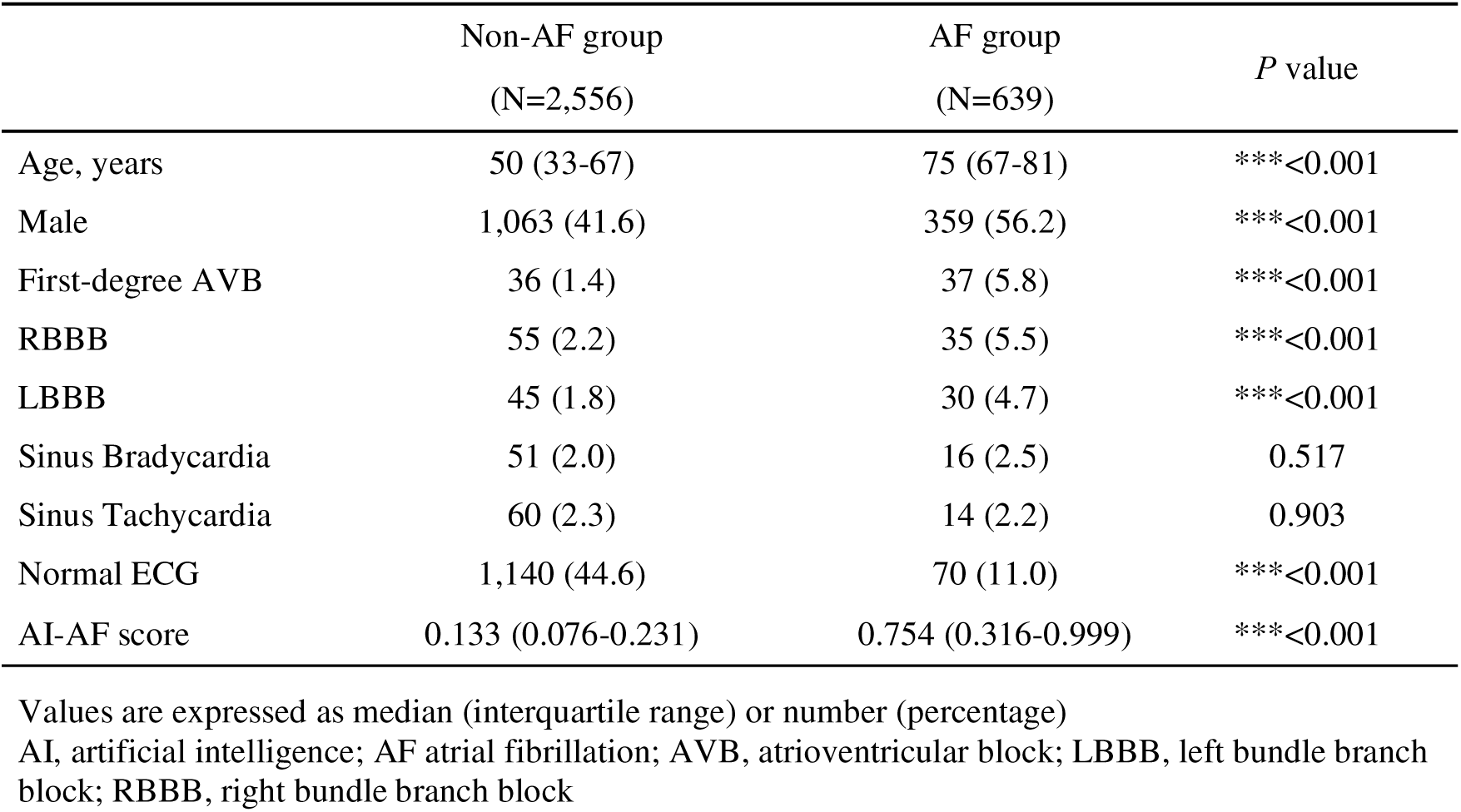
Description of external validation dataset (CODE 15%)

The AI model effectively distinguished AF patients, achieving an AUC-ROC of 0.884 (95% CI: 0.869-0.900), with sensitivity of 74.3% (70.8-77.7%), and specificity of 86.5% (85.2-87.8%). The PLR and NLR were 5.52 (4.96-6.16) and 0.30 (0.26-0.34), respectively (Table 4). After adjusting for age and sex, the AUC-ROC improved to 0.906 (95% CI: 0.893-0.919) (Figure 3), with corresponding changes in sensitivity and specificity to 82.9% (79.8-85.8%) and 82.2% (80.7-83.7%), respectively.

**Figure 3.**
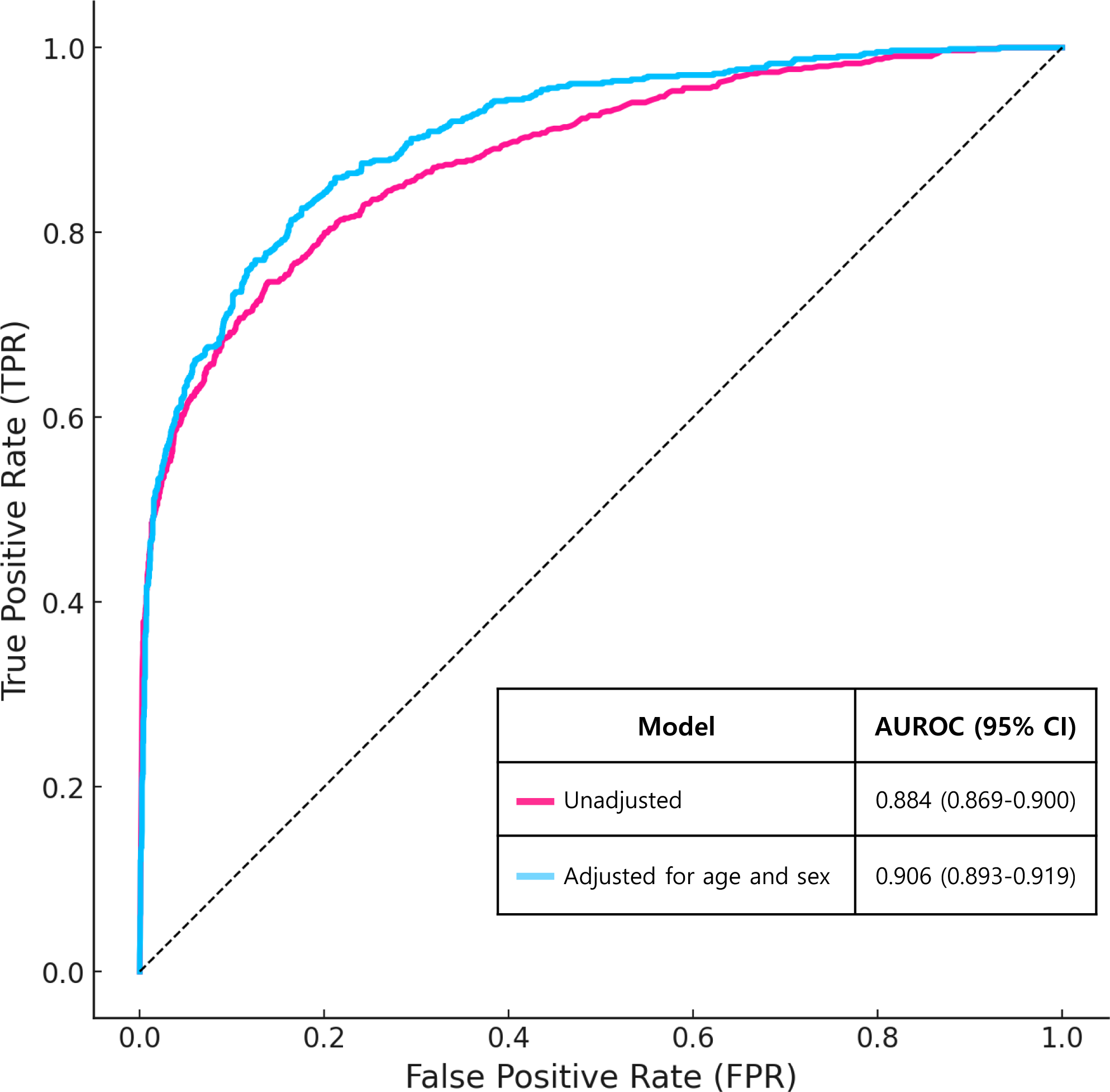
Receiver operating characteristics curve analysis for EBC’s AF prediction

**Table 4.**
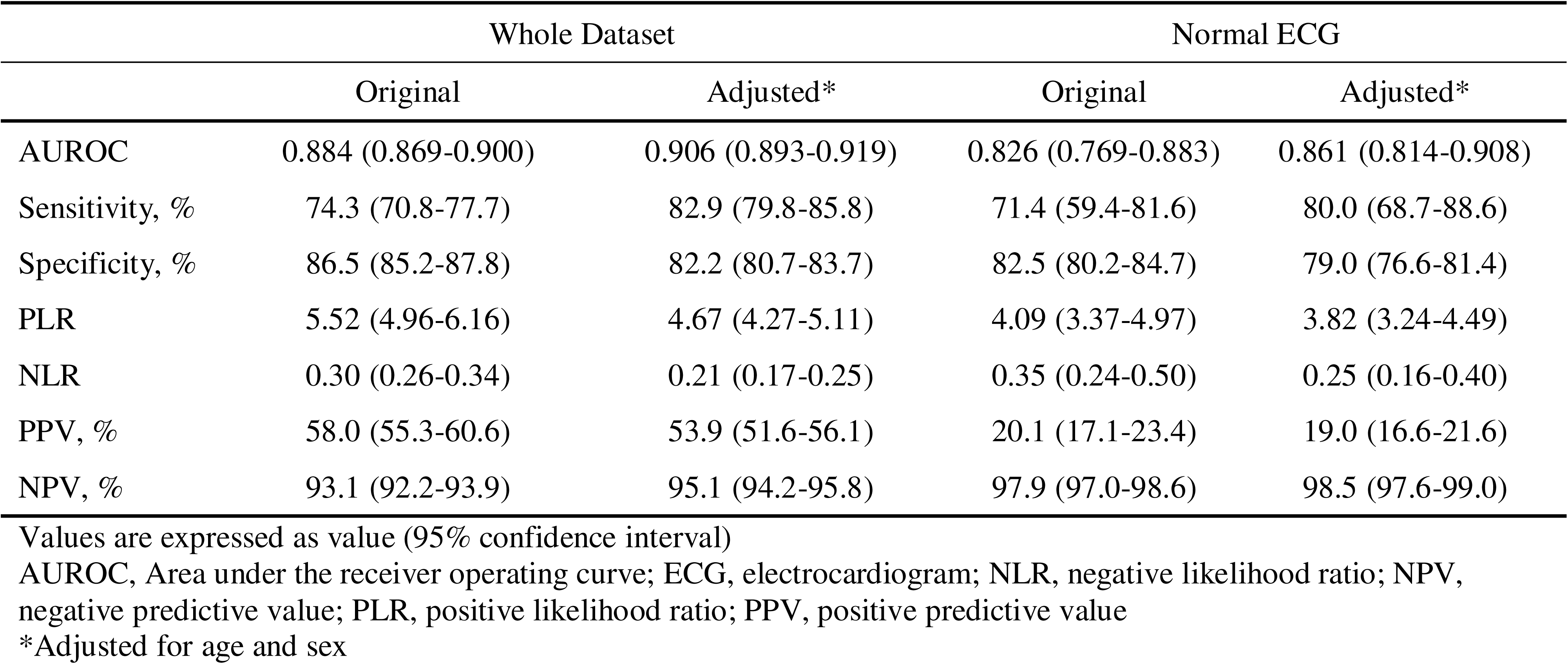
Performance evaluation in the external validation dataset.

The adjusted PLR and NLR were 4.67 (4.27-5.11) and 0.21 (0.17-0.25), respectively (Table 4). In the subgroup analysis of patients with "Normal ECG" interpretations, the unadjusted AUC-ROC, sensitivity, and specificity were 0.826 (0.769-0.883), 71.4% (59.4-81.6%), and 82.5% (80.2-84.7%), respectively. After adjusting for age and sex, these values improved to 0.861 (0.814-0.908), 80.0% (68.7-88.6%), and 79.0% (76.6-81.4%), respectively (Table 4).

## Discussion

This study represents a substantial advancement in applying AI to cardiology, particularly in predicting concurrent or incident AF using 12-lead SR ECG images. Our AI model demonstrated high predictive accuracy in internal validation, achieving an AUROC of 0.907. Furthermore, in external interethnic validation using the CODE 15% data, the model exhibited robust performance with an AUROC of 0.884, which further improved to 0.906 after adjusting for age and sex. The model’s effectiveness in the "Normal ECG" subgroup highlights its potential to be used in the general healthy population to screen individuals at high risk of developing AF.

A key advantage of this model over previous approaches in AI-enabled AF screenings is its ability to process ECG images, whereas conventional methods have predominantly relied on one-dimensional raw ECG signals.^4–6^ Our approach better aligns in terms of clinical practices and enhances the model’s practical applicability as ECGs are commonly stored and interpreted in printed format.^14^ For example, the model can be seamlessly integrated into telemedicine system using smartphone devices or scanners. In the current landscape of digital health, the model’s compatibility with ECG images allows for remote monitoring and telemedicine applications. This improved versatility, while maintaining predictability comparable to previously published models,^4–6^is one of the key strengths of our AI algorithm.

Demonstration of consistent accuracy in the interethnic validation is another noteworthy finding of this study, as ECG configurations can vary slightly across different ethnic group. For instance, QRS voltages have been reported to be higher in young Black males compared to other ethnicities, including Caucasian, East Asian, and Indian populations.^15^ And PR interval appears to be slightly longer in Asians compared with Whites.^16^ Since High QRS voltages and prolonged PR intervals are well-known ECG findings which are potentially related to AF, due to their links with diastolic dysfunction and atrial fibrosis.^17,18^ The model’s ability to maintain high accuracy across the different ethnicities suggests that it is impervious to ethnic ECG variations.

The model’s capability to accurately identify high-risk individuals for AF could have a significant impact on clinical practice. Although there is currently no strong evidence to support the benefits of AF screening in general population,^19^ this model could be particularly useful in high-risk groups such as patients with embolic stroke undetermined source (ESUS). Many researchers have identified ESUS patients as an optimal population for AI-based AF screening, given that their high AF prevalence and the clinical consequence of missed AF diagnoses.^4–6,20,21^ A previous version of our model demonstrated excellent performance in identifying AF in ESUS patients.^20^ By integrating this AI model with traditional AF risk factors, such as age, left atrial diameter and burden of atrial ectopic beats, its predictability can be further enhanced, providing valuable insights for clinical decision-making and downstream management strategies.^20^

Several limitations should be addressed. First, the external validation was conducted retrospectively. Although the dataset included a large number of patients, prospective validation is still necessary to further assess the model’s performance. Second, only a limited number of variables were available for multivariable analysis, as detailed patient-level information was lacking in the external validation dataset. Third, the definition of “Normal ECG” in the external validation dataset was not explicitly detailed in the dataset instruction. We carefully assume that it referred to sinus rhythm with a normal range of heart rate (60-100bpm) without significant rhythm or conduction disorder.

In conclusion, this study presents an innovative AI model for identifying AF based on sinus rhythm ECGs. The model has improved accessibility by leveraging AI imaging analysis techniques and has demonstrated strong performance across different ethnicities, comparable to previously published studies. We believe that this model has the potential to play a valuable role in identifying current or incident AF, alongside conventional risk factors, on a global scale.

## Data Availability

All data produced in the present study are available upon reasonable request to the authors

## ACKNOWLEDGEMENTS

This research was supported by a grant of the Korea Health Technology R&D Project through the Korea Health Industry Development Institute (KHIDI), funded by the Ministry of Health & Welfare, Republic of Korea (grant number: RS-2023-00265933), and the Technological Innovation R&D Program (SCALEUP TIPS), funded by the Ministry of SMEs and Startups (grant number: RS-2024-00415492).

## Disclosure

Joonghee Kim developed the algorithm and is the founder and CEO of the startup company, ARPI Inc. Youngjin Cho has worked at ARPI Inc. as a research director. The remaining authors declare no conflicts of interest

## Author Contributions

Conceptualization: Lee JH, Kim J, Oh IY, Cho Y. Data curation: Kim J. Formal analysis: Kim J. Funding acquisition: Kim J. Investigation: Lee JH, Kim J. Cho J. Methodology: Lee JH, Kim J, Cho Y. Project administration: Kim J, Cho Y. Resources: Kim J, Cho Y. Software: Kim J, Cho Y. Supervision: Cho Y. Validation: Kim J, Cho Y, Lee JH. Visualization: Kim J, Lee JH. Writing – original draft: Lee JH. Writing – review & editing: Choi YY, Cho J, Oh IY

---

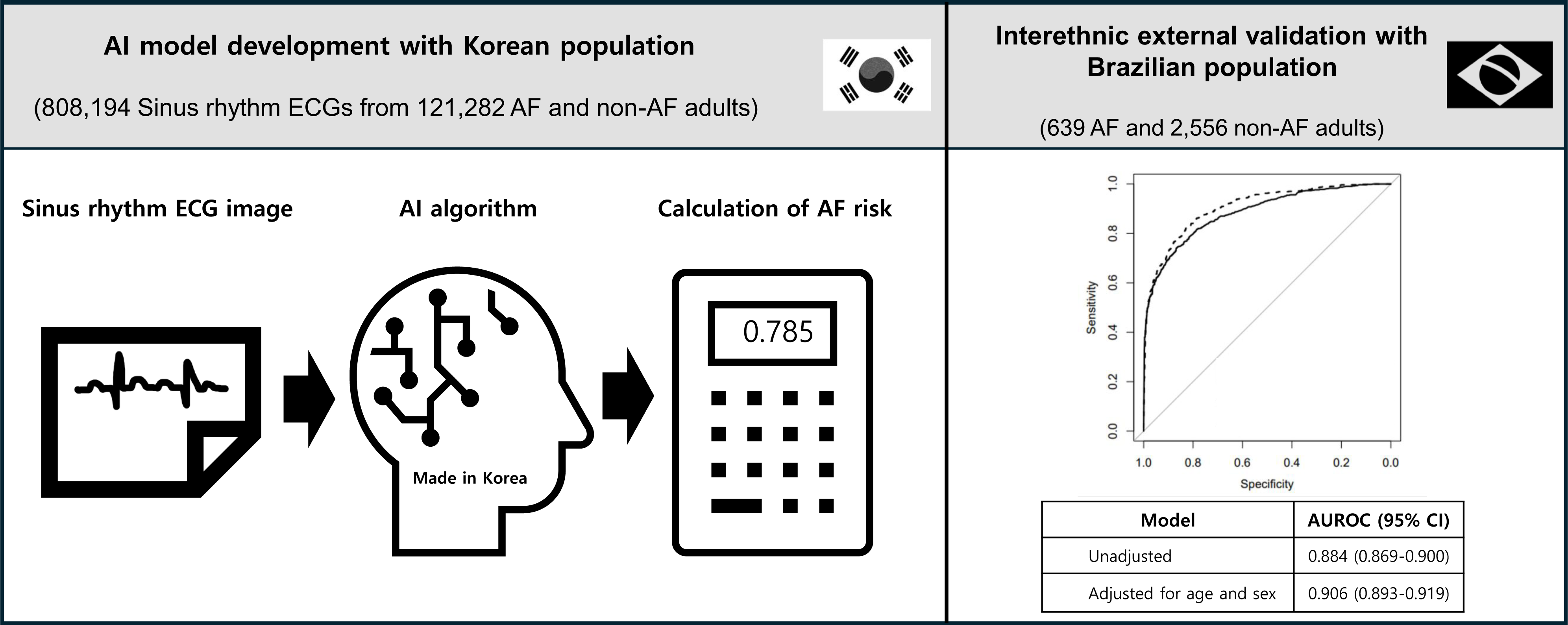

## Notes

### Author Declarations

This study was approved by the Institutional Review Board of Seoul National University Bundang Hospital (IRB No.: B-2205-757-002), and informed consent was waived due to the retrospective study design.

