## Supplementary material for "Interethnic Validation of Artificial Intelligence for prediction of Atrial Fibrillation Using Sinus Rhythm Electrocardiogram": Tables

| Table 1. Description of the development dataset. | | | | | | |
| --- | --- | --- | --- | --- | --- | --- |
|  | Training dataset | | | Internal validation dataset | | |
|  | Non-AF | AF | *P* | Non-AF | AF | *P* |
|  | (N=442,931) | (N=284,531) |  | (N=9,144) | (N=1,209) |  |
| Age, years | 63 (53-72) | 72 (63-79) | ***<0.001 | 57 (47-67) | 70 (60-78) | ***<0.001 |
| Male | 248,287 (56.1) | 177,568 (62.4) | ***<0.001 | 4,681 (51.2) | 775 (64.1) | ***<0.001 |
| Health Checkup or   Outpatient Setting | 324,920 (73.4) | 140,194 (49.3) | ***<0.001 | 6,859 (75.0) | 410 (33.9) | ***<0.001 |
| Normal ECG Reading | 143,736 (32.5) | 17,092 (6.0) | ***<0.001 | 3,806 (41.6) | 181 (15.0) | ***<0.001 |
| Sinus Rhythm* | 405,204 (91.5) | 108,721 (38.2) | ***<0.001 | 9,144 (100.0) | 1,209 (100.0) | - |
| * Sinus Rhythm with 50≤HR<100 Values are expressed median (interquartile range) or number (percentage) ECG, electrocardiogram; AF, atrial fibrillation | | | | | | |

| Table 2. Internal validation performance. | | | | | | | |
| --- | --- | --- | --- | --- | --- | --- | --- |
|  | AUROC | Sensitivity, % | Specificity, % | PPV, % | NPV, % | PLR | NLR |
| ALL | 0.907 (0.897-0.916) | 80.6 (78.3-82.8) | 85.0 (84.2-85.7) | 41.2 (39.9-42.6) | 97.10 (96.76-97.41) | 5.36 (5.07-5.67) | 0.23 (0.20-0.26) |
| Health Checkup or Outpatient Setting | 0.874 (0.856-0.891) | 70.5 (65.8-74.8) | 88.8 (88.0-89.5) | 27.4 (25.6-29.2) | 98.1 (97.7-98.3) | 6.29 (5.74-6.89) | 0.33 (0.29-0.39) |
| Normal ECG | 0.852 (0.824-0.880) | 76.1 (69.3-82.1) | 76.8 (75.4-78.2) | 14.1 (12.8-15.2) | 98.5 (98.0-98.8) | 3.28 (2.97-3.63) | 0.31 (0.24-0.40) |
| Values are expressed as value (95% confidence interval)  AUROC, Area under the receiver operating curve; ECG, electrocardiogram; NLR, negative likelihood ratio; NPV, negative predictive value; PLR, positive likelihood ratio; PPV, positive predictive value | | | | | | | |

| Table 3. Description of external validation dataset (CODE 15%) | | | |
| --- | --- | --- | --- |
|  | Non-AF group | AF group | *P* value |
|  | (N=2,556) | (N=639) |  |
| Age, years | 50 (33-67) | 75 (67-81) | ***<0.001 |
| Male | 1,063 (41.6) | 359 (56.2) | ***<0.001 |
| First-degree AVB | 36 (1.4) | 37 (5.8) | ***<0.001 |
| RBBB | 55 (2.2) | 35 (5.5) | ***<0.001 |
| LBBB | 45 (1.8) | 30 (4.7) | ***<0.001 |
| Sinus Bradycardia | 51 (2.0) | 16 (2.5) | 0.517 |
| Sinus Tachycardia | 60 (2.3) | 14 (2.2) | 0.903 |
| Normal ECG | 1,140 (44.6) | 70 (11.0) | ***<0.001 |
| AI-AF score | 0.133 (0.076-0.231) | 0.754 (0.316-0.999) | ***<0.001 |
| Values are expressed as median (interquartile range) or number (percentage) AI, artificial intelligence; AF atrial fibrillation; AVB, atrioventricular block; LBBB, left bundle branch block; RBBB, right bundle branch block | | | |

| Table 4. Performance evaluation in the external validation dataset | | | | |
| --- | --- | --- | --- | --- |
|  | Whole Dataset | | Normal ECG | |
|  | Original | Adjusted* | Original | Adjusted* |
| AUROC | 0.884 (0.869-0.900) | 0.906 (0.893-0.919) | 0.826 (0.769-0.883) | 0.861 (0.814-0.908) |
| Sensitivity, % | 74.3 (70.8-77.7) | 82.9 (79.8-85.8) | 71.4 (59.4-81.6) | 80.0 (68.7-88.6) |
| Specificity, % | 86.5 (85.2-87.8) | 82.2 (80.7-83.7) | 82.5 (80.2-84.7) | 79.0 (76.6-81.4) |
| PLR | 5.52 (4.96-6.16) | 4.67 (4.27-5.11) | 4.09 (3.37-4.97) | 3.82 (3.24-4.49) |
| NLR | 0.30 (0.26-0.34) | 0.21 (0.17-0.25) | 0.35 (0.24-0.50) | 0.25 (0.16-0.40) |
| PPV, % | 58.0 (55.3-60.6) | 53.9 (51.6-56.1) | 20.1 (17.1-23.4) | 19.0 (16.6-21.6) |
| NPV, % | 93.1 (92.2-93.9) | 95.1 (94.2-95.8) | 97.9 (97.0-98.6) | 98.5 (97.6-99.0) |
| Values are expressed as value (95% confidence interval)  AUROC, Area under the receiver operating curve; ECG, electrocardiogram; NLR, negative likelihood ratio; NPV, negative predictive value; PLR, positive likelihood ratio; PPV, positive predictive value  *Adjusted for age and sex | | | | |
